# Rare variants in *IFFO1, DTNB* and *NLRC3* associate with Alzheimer’s disease CSF profile of neuronal injury and inflammation

**DOI:** 10.1101/2021.07.10.21260177

**Authors:** Alexander Neumann, Fahri Küçükali, Isabelle Bos, Stephanie J.B. Vos, Sebastiaan Engelborghs, Tim De Pooter, Geert Joris, Peter De Rijk, Ellen De Roeck, Magda Tsolaki, Frans Verhey, Pablo Martinez-Lage, Mikel Tainta, Giovanni Frisoni, Oliver Blin, Jill Richardson, Régis Bordet, Philip Scheltens, Julius Popp, Gwendoline Peyratout, Peter Johannsen, Lutz Frölich, Rik Vandenberghe, Yvonne Freund-Levi, Johannes Streffer, Simon Lovestone, Cristina Legido-Quigley, Mara ten Kate, Frederik Barkhof, Mojca Strazisar, Henrik Zetterberg, Lars Bertram, Pieter Jelle Visser, Christine van Broeckhoven, Kristel Sleegers, EMIF-AD study group, Alzheimer’s Disease Neuroimaging Initiative

## Abstract

Alzheimer’s disease (AD) biomarkers represent several neurodegenerative processes, such as synaptic dysfunction, neuronal inflammation and injury, as well as amyloid pathology. We performed an exome-wide rare variant analysis of six AD biomarkers (β-amyloid, total/phosphorylated tau, Nfl, YKL-40, and Neurogranin) to discover genes associated with these markers. Genetic and biomarker information was available for 480 participants from two studies: EMIF-AD and ADNI. We applied a principal component (PC) analysis to derive biomarkers combinations, which represent statistically independent biological processes. We then tested whether rare variants in 9,576 protein-coding genes associate with these PCs using a Meta-SKAT test. We also tested whether the PCs are intermediary to gene effects on AD symptoms with a SMUT test. One PC loaded on Nfl and YKL-40, indicators of neuronal injury and inflammation. Three genes were associated with this PC: *IFFO1, DTNB* and *NLRC3*. Mediation tests suggest, that these genes also affect dementia symptoms via inflammation/injury. We also observed an association between a PC loading on Neurogranin, a marker for synaptic functioning, with *GABBR2* and *CASZ1*, but no mediation effects. The results suggest that rare variants in *IFFO1, DTNB* and *NLRC3* heighten susceptibility to neuronal injury and inflammation, potentially by altering cytoskeleton structure and immune activity disinhibition, resulting in an elevated dementia risk. *GABBR2* and *CASZ1* were associated with synaptic functioning, but mediation analyses suggest that the effect of these two genes on synaptic functioning is not consequential for AD development.

## Introduction

Alzheimer’s disease (AD) is a neurodegenerative disease with an estimated heritability of 63%[1], with common variants explaining 9-31% of disease liability.[2] Several studies have also found a contribution of rare variants in genes such as *TREM2* and *ABCA7* towards AD.[3–5] A popular approach to discover AD relevant rare variants is the use of whole-exome/genome sequencing to assess rare-variants globally, and then to associate each variant with AD status.[6] However, the occurrence of AD is caused by a combination of pathways involving inflammation, cholesterol metabolism, tau pathology, endosome or ubiquitin-related functioning.[7] Individuals with the same symptoms can differ regarding the pathways contributing to their symptoms. At the same time, different AD relevant genes may act on different pathways. Studying AD status as an outcome may therefore mask genetic effects, which only affect specific pathways or patients subsets. In this study we focus on six CSF biomarkers, which reflect different AD relevant disease processes. The examined biomarkers are amyloid beta peptide 42 (Aβ), tau, phosphorylated tau (pTau), neurofilament light chain (Nfl), YKL-40 and Neurogranin (Ng). The application of these proteins/peptides has been reviewed previously[8, 9] and is summarized below:

Aβ and tau are the two most well established AD biomarkers that reflect the defining neuropathological hallmarks of AD (amyloid plaques and tau tangles).[8, 9] Plaque deposits of Aβ in extracellular space are one of the earliest disease processes, with accumulation beginning many years before first symptoms emerge.[10] Aβ CSF levels are inversely related to brain levels, *i*.*e*., lower levels of the 42 amino acid-long aggregation-prone Aβ (Aβ42) in CSF are indicative of higher brain deposition of the protein.[11] pTau is one of the components of neurofibrillary tangles. Both total and pTau are increased in the CSF of AD patients.[11] Compared to Aβ, it is a more concurrent state marker of neurodegeneration, with elevated levels occurring later during disease progression.[9] Nfl is a building block of axons and higher levels in CSF are indicative of neuronal injury. [12] Levels of Nfl are elevated in AD patients, but it is not a specific marker of AD. [9] Another non-specific biomarker is YKL40, which represents astrocytic activation and neuronal inflammation. YKL-40 is associated with AD status and other neuropathologies. [11, 13] Finally, neurogranin is a postsynaptic protein related to synaptic functioning, cognition and plasticity. Importantly, levels are higher in AD patients and other dementias. [14]

Most genome-wide analyses use a case-control design, but some have also examined CSF and plasma biomarkers. A recent genome-wide association study (GWAS) has examined common variation in relation to Aβ and tau levels in CSF, identifying novel associations between *ZFHX3* and CSF-Aβ38 and Aβ40 levels, and confirmed a previously described sex-specific association between SNPs in *GMNC* and CSF-tTau.[15] A more recent GWAS on these datasets further identified common-variant associations between *TMEM106B* and CSF-Nfl and *CPOX* and CSF-YKL-40.[16] Another study investigated rare variants underlying plasma Aβ using whole-exome sequencing, identifying several exome-wide significant genes.[17]

With this study, we took a pathway approach by analyzing rare variants in relation to distinct AD-related pathologic processes reflected by six different CSF biomarkers. As we are mainly interested in the genetics of the underlying disease processes, which can be represented by multiple biomarkers, as opposed to the biology of single biomarkers per se, we apply a multivariate approach to analyze multiple biomarkers jointly. Specifically, we applied a principal component analysis (PCA) to identify independent clusters of biomarkers representing different biological processes. A PCA approach is not only conceptually appealing, but may also improve power.[18, 19]

To further improve power and generalizability, we performed a mega-analysis of two multi-center studies: the European Medical Information Framework for Alzheimer’s Disease Multimodal Biomarker Discovery (EMIF-AD MBD) study[20] and the Alzheimer’s Disease Neuroimaging Initiative (ADNI)[21].

## Methods

### Participants

This study was embedded in the EMIF-AD MBD project, a consortium of European cohort studies with the aim to increase understanding of AD pathophysiology and discover diagnostic and prognostic biomarkers.[20] The EMIF-AD MBD study includes participants with no cognitive impairment, mild cognitive impairment (MCI) or AD. Extensive phenotype information is available on diagnosis, cognition, CSF and imaging biomarkers. Genetic assessments include genome-wide SNP and DNA methylation array data, as well as whole-exome sequencing. Written informed consent for use of data, samples and scans was obtained from all participants before inclusion in EMIF-AD MBD. The medical ethics committee at each site approved the study.[20]

We further included ADNI to increase power and generalizability.[21] Data used in the preparation of this article were obtained from adni.loni.usc.edu. ADNI was launched in 2003 as a public-private partnership, led by Principal Investigator Michael W. Weiner, MD. The primary goal of ADNI has been to test whether serial magnetic resonance imaging (MRI), positron emission tomography (PET), other biological markers, and clinical and neuropsychological assessment can be combined to measure the progression of MCI and early AD.

For the main analysis we selected participants, who were assessed with exome-wide sequencing, had no known pathogenic mutations, were unrelated and had information on at most one CSF biomarker missing, resulting in a total sample size of 480 participants. Participants had mostly European ancestry (98.8%). Primary analyses were based on this multi-ancestry sample, but European ancestry only analyses are provided as sensitivity analysis (Supplementary Methods).

### Measures

#### Genotyping

Whole exome-sequencing in EMIF was performed using an Illumina NextSeq500 platform using paired-end reads on DNA samples hybridized with SeqCap EZ Human Exome Kit v3.0 (Roche). In ADNI whole exome-sequencing was performed using the Illumina HiSeq2000 platform on DNA samples hybridized with the Agilent’s SureSelect Human All Exon 50 Mb kit.[22] The same quality control pipeline was then applied to both studies (Supplementary Methods). Post-analysis, we retained only genes with at least two rare variant carriers in each study to reduce Type-1 error.

#### CSF Biomarkers and Dementia Symptoms

CSF has been obtained via lumbar puncture and biomarker levels analyzed as previously described.[21, 23] In brief, in EMIF the V-PLEX Plus AbPeptidePanel 1 Kit assessed Aβ and INNOTEST ELISA was used for tau.[23] In ADNI the Elecsys CSF immunoassay with a cobas e 601 analyzer was used to measure Aβ and tau concentration.[24] In both EMIF and ADNI Nfl was analyzed using ELISA.[23, 25] Ng was assessed using an immunoassay in EMIF[23] and electrochemiluminescence technology in ADNI[26]. YKL-40 was measured with an ELISA kit in EMIF[23] and LC/MRM‐MS proteomics in ADNI[27]. As the ADNI proteomics data contained two peptide sequences, with two ion frequencies each, we averaged across these four values after z-score standardization. Both studies used the Mini-Mental State Examination, a 30 item questionnaire to assess dementia symptoms.[28]

### Statistical analysis

#### PCA

We first performed a PCA across both studies to identify and compute independent components using linear combinations of the measured biomarkers. Biomarkers showed extreme skewness, which can distort findings.[29] We therefore transformed all biomarkers with rank based inverse normal transformation (INT) within each cohort. The resulting z-score also harmonizes the scale between the cohorts. We used a PCA-based imputation approach, as implemented in missMDA, to account for missing levels of biomarkers.[30] To determine the optimum number of dimensions, we applied leave-one-out cross-validation minimizing the squared error of prediction. The PCA was performed in the same analysis sample as the main genetic analysis, but see sensitivity analyses for results in a larger sample not restricted by genetic information (n=1158). PCA scores were computed with the psych package.[31] All analyses were performed in R 4.0.3.[32]

#### Gene-based tests

We focused on rare variants with potentially large impacts on pathogenic processes. We analyzed rare protein-coding variants with a minor allele frequency below 1% in the EMIF/ADNI population and associated them with biomarker PCs. In secondary analyses, we further prioritized loss-of-function variants.

We used a SKAT-O test,[33] a kernel-based method, as implemented in MetaSKAT, allowing for heterogeneous effects between studies. MetaSKAT is an extension of the original SKAT test designed for meta-analyses.[34] As individual level data was available for both studies, we performed a mega-analysis on combined datasets.

All analyses were adjusted for sex, age and genetic ancestry. In EMIF we used the first four genome-wide PCs and in ADNI the first ten, taking into account the higher population admixture. We performed analyses both with and without adjusting for diagnosis (dummy coding for MCI and AD), to avoid collider bias in case the genetic variant and PC are both independently causative of AD.[35] To characterize which specific variants drive the gene associations, we followed up gene hits with a single variant regression analysis model analogous to the SKAT analyses.

#### Mediation tests

Genes with exome-wide significance were followed up with mediation tests. The mediation models tested whether genes impact dementia via their influence on the examined neurodegenerative process. The outcome in the models were MMSE scores and the mediator was the PC showing an exome-wide significant association with the gene. MMSE scores were normalized using a previously described method.[36] Mediation tests were performed with SMUT, an intersection-union test based on SKAT.[37] We regressed outcomes on sex, age and genetic ancestry and z-score standardized the resulting residuals within cohorts. Normalized MMSE scores were residualized jointly across cohorts using sex, age and genetic ancestry. These residuals were also used to correlate the PCs with MMSE to better characterize the PCs using spearman correlations. We also looked up the total effect of the genes with MMSE using the same MetaSKAT model as used with the PCs.

### Data availability

To comply with EU law and participant privacy, individual-level clinical data cannot be shared publicly, but can be requested via EMIF-AD (https://emif-catalogue.eu;http://www.emif.eu/about/emif-ad).

## Results

### Demographics

Descriptive statistics are presented in Table 1. Both EMIF and ADNI represent an elderly population of comparable ages, but EMIF recruited a larger proportion of participants with AD. The included sample of ADNI concerned only participants with no or mild cognitive impairment, resulting in a higher mean score of the MMSE, indicating better performance.

**Table 1:**
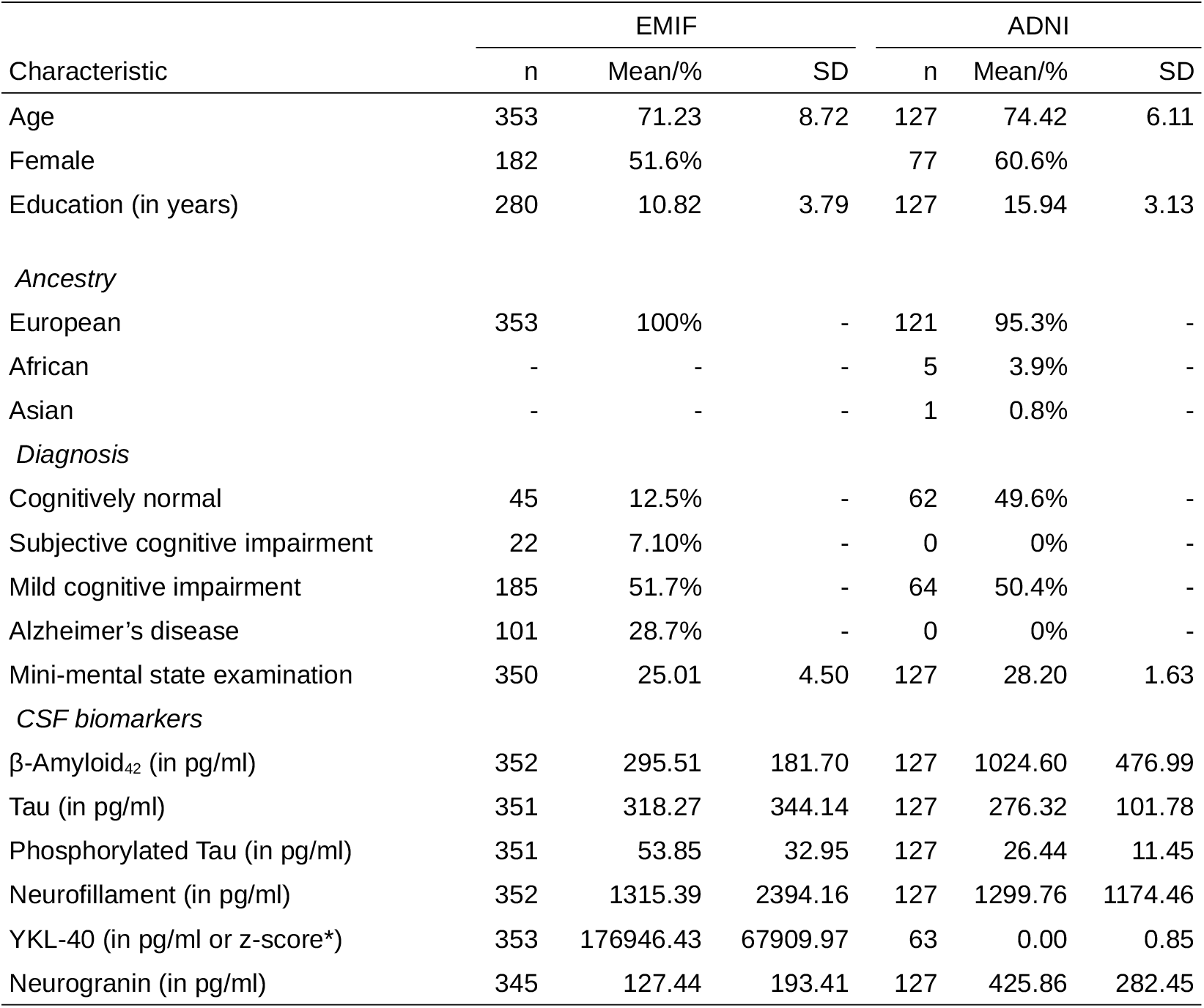
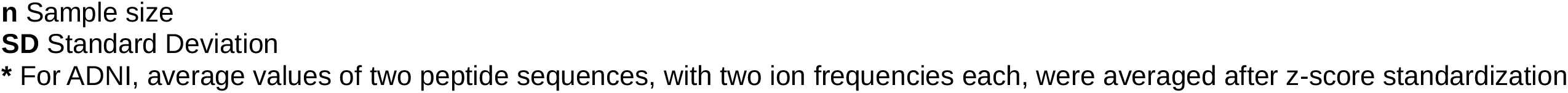
Participant characteristics. Demographic information and descriptive statistics.

### PCA

The cross-validation informed the use of five components (Table 2). The first component loaded strongly on the tau measures and moderately on Ng and YKL-40. Given the component’s strong loading on tTau and pTau, but also at the same time moderate loading on other aspects of neurodegeneration, we interpret the component as representing tau pathology and neurodegeneration in general. This tau pathology/degeneration PC was negatively correlated with MMSE (r=-0.19). The second PC loaded mostly on Nfl, with a moderate loading on YKL-40, thus we can interpret it as indicating neuronal injury and inflammation. This PC correlated with MMSE negatively as well (r=-0.22). The third component was highly specific to Aβ and had the strongest correlation with dementia symptoms in the expected positive direction (r=0.28). The fourth component loaded mostly on YKL-40 with weak loadings on tTau and Nfl. This component did not correlate with MMSE scores and was therefore labeled Non-AD Inflammation. The final component loaded mostly on Ng, with weak loadings on the tau measures, but again no correlation with dementia symptoms and therefore is named Non-AD Synaptic functioning

**Table 2:** PCA results. Principal component analysis of CSF biomarkers per study (EMIF/ADNI) and across studies (Mega). Component loadings of each biomarker (first column) on five principal components (column groups two to six) are displayed in order of the component’s explained variance (R^2^). Analyses are based on 480 participants (EMIF: 354, ADNI: 127).

| | Tau pathology/<br>Degeneration | | | Injury/<br>Inflammation | | | Aβ Pathology | | | Non-AD<br>Inflammation | | | Non-AD<br>Synaptic functioning | | |
| --- | --- | --- | --- | --- | --- | --- | --- | --- | --- | --- | --- | --- | --- | --- | --- |
|  | EMIF | ADNI | Mega | EMIF | ADNI | Mega | EMIF | ADNI | Mega | EMIF | ADNI | Mega | EMIF | ADNI | Mega |
| *Loadings* |  |  |  |  |  |  |  |  |  |  |  |  |  |  |  |
| Tau | 0.84 | 0.91 | 0.86 | 0.25 | 0.19 | 0.23 | -0.06 | -0.04 | -0.05 | 0.28 | 0.23 | 0.27 | 0.27 | 0.26 | 0.28 |
| pTau | 0.91 | 0.93 | 0.92 | 0.10 | 0.14 | 0.11 | -0.05 | -0.14 | -0.07 | 0.16 | 0.20 | 0.17 | 0.26 | 0.23 | 0.26 |
| Aβ | -0.06 | -0.09 | -0.07 | -0.05 | -0.04 | -0.04 | 1.00 | 1.00 | 1.00 | -0.05 | -0.01 | -0.04 | 0.03 | 0.02 | 0.03 |
| NFL | 0.19 | 0.19 | 0.19 | 0.94 | 0.95 | 0.94 | -0.05 | -0.04 | -0.05 | 0.25 | 0.25 | 0.25 | 0.10 | 0.07 | 0.09 |
| YKL-40 | 0.31 | 0.28 | 0.31 | 0.31 | 0.28 | 0.30 | -0.06 | -0.01 | -0.05 | 0.88 | 0.91 | 0.89 | 0.18 | 0.10 | 0.16 |
| Ng | 0.45 | 0.57 | 0.48 | 0.12 | 0.09 | 0.11 | 0.05 | 0.04 | 0.05 | 0.19 | 0.13 | 0.18 | 0.86 | 0.80 | 0.85 |
| R<sup>2</sup> | 0.32 | 0.36 | 0.33 | 0.18 | 0.17 | 0.18 | 0.17 | 0.17 | 0.17 | 0.17 | 0.17 | 0.17 | 0.16 | 0.13 | 0.15 |
| MMSE correlation | -0.21 | -0.15 | -0.19 | -0.25 | -0.15 | -0.22 | 0.33 | 0.13 | 0.28 | -0.07 | -0.01 | -0.05 | -0.02 | 0.01 | -0.02 |

**R<sup>2</sup>** Variance explained by component
**MMSE correlation** Spearman correlation between principal component and MMSE scores. MMSE scores were residualized for age, sex and genetic ancestry.

The PCA results were highly consistent across both studies, however, association magnitudes with MMSE differed between studies, with ADNI showing lower effect sizes. We also performed a sensitivity analysis in a larger sample not filtered for availability of genetic assessments. The same five components were identified and loadings were nearly identical, differing at most by .05 (Supplementary Table 1).

### Whole-exome rare variant analysis: Protein-coding variants

#### Quality Control

After QC, 9,576 genes remained with at least four rare variant carriers. Exome-wide significance was therefore set at p=5.2*10^−6^ (Bonferroni correction). Lambda was 1 or lower, suggesting that test statistics were not inflated due to population stratification or wide-spread collider bias (Supplementary Figure 1).

#### No diagnosis adjustment

No gene passed exome-wide significance for the Tau pathology/Degeneration, Aβ Pathology, or Non-AD synaptic functioning PC. See Table 3 for gene-based results, Figure 1 for Manhattan plots, Figure 2 for outcome distributions and Supplementary Table 2 for single-variant results.

**Table 3:**
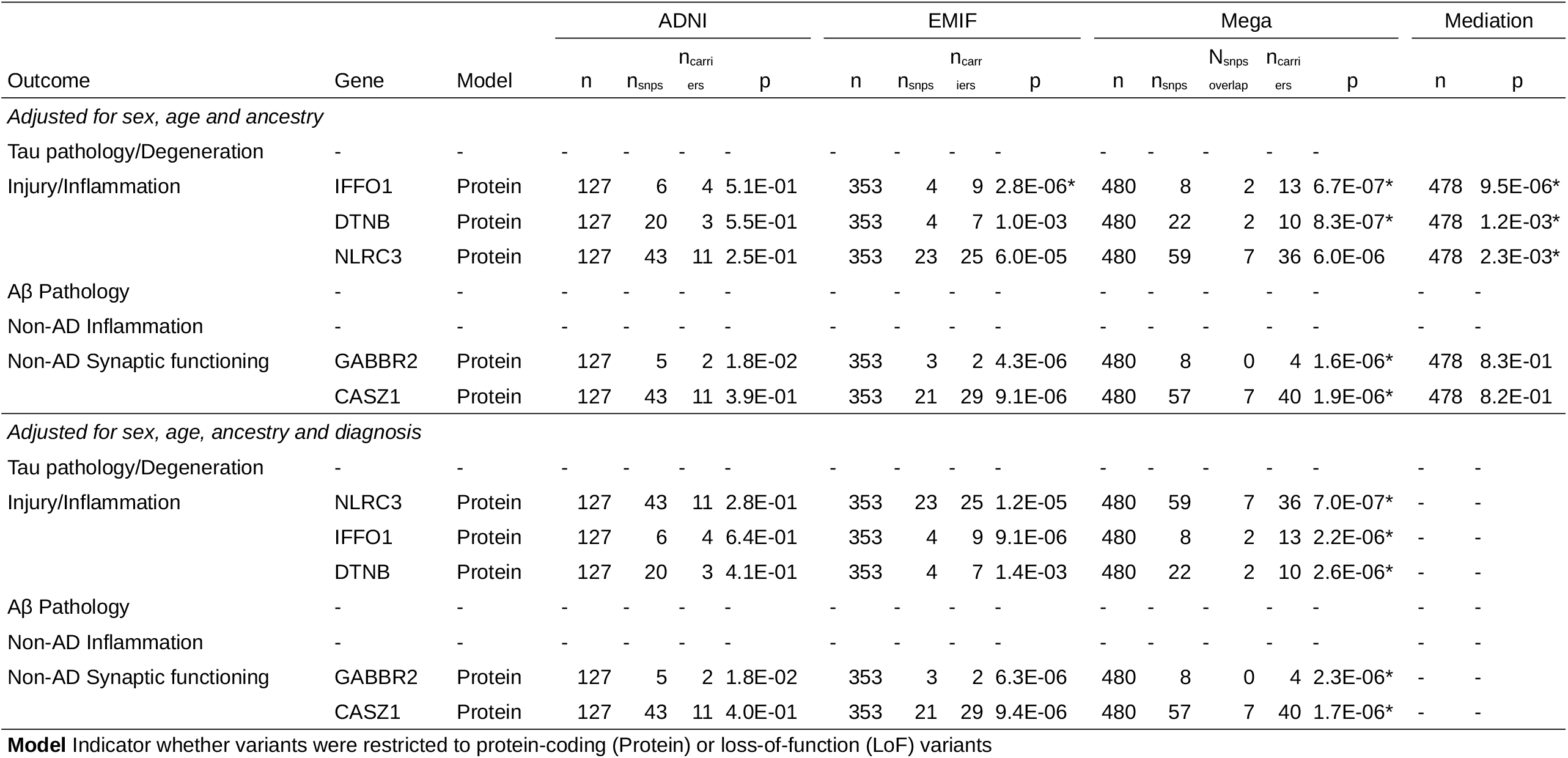
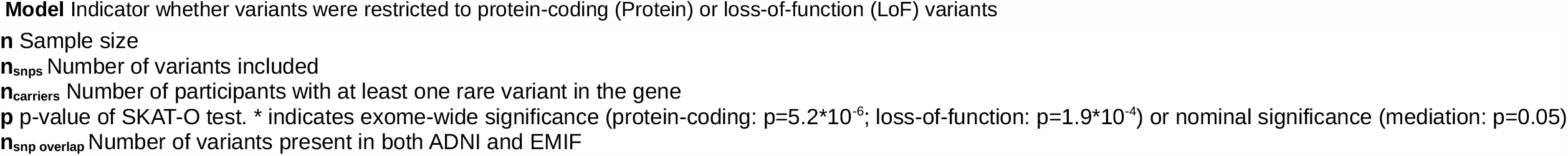
Genes with exome-wide significant associations. Results for exome-wide rare-variant and mediation analyses. Rare (MAF<1%) protein-coding variants in 13,363 genes were tested on a gene level in the protein-coding model and 536 genes in the loss-of-function model. Each gene was associated with five principal component scores of CSF biomarkers, representing different neurodegenerative processes. P-values (p) were obtained from gene-based SKAT-O tests. SMUT tested mediation on dementia symptoms (MMSE scores) via changes in the principal components. All tests were adjusted for sex, age and genetic ancestry (top group). In separate models, we additionally adjusted for diagnosis status (bottom group). Only genes with exome-wide significant association in the mega-analysis (Mega) are displayed (protein-coding: p<5.2*10^−6^; lof: p<1.9*10^−4^).

**Figure 1:**
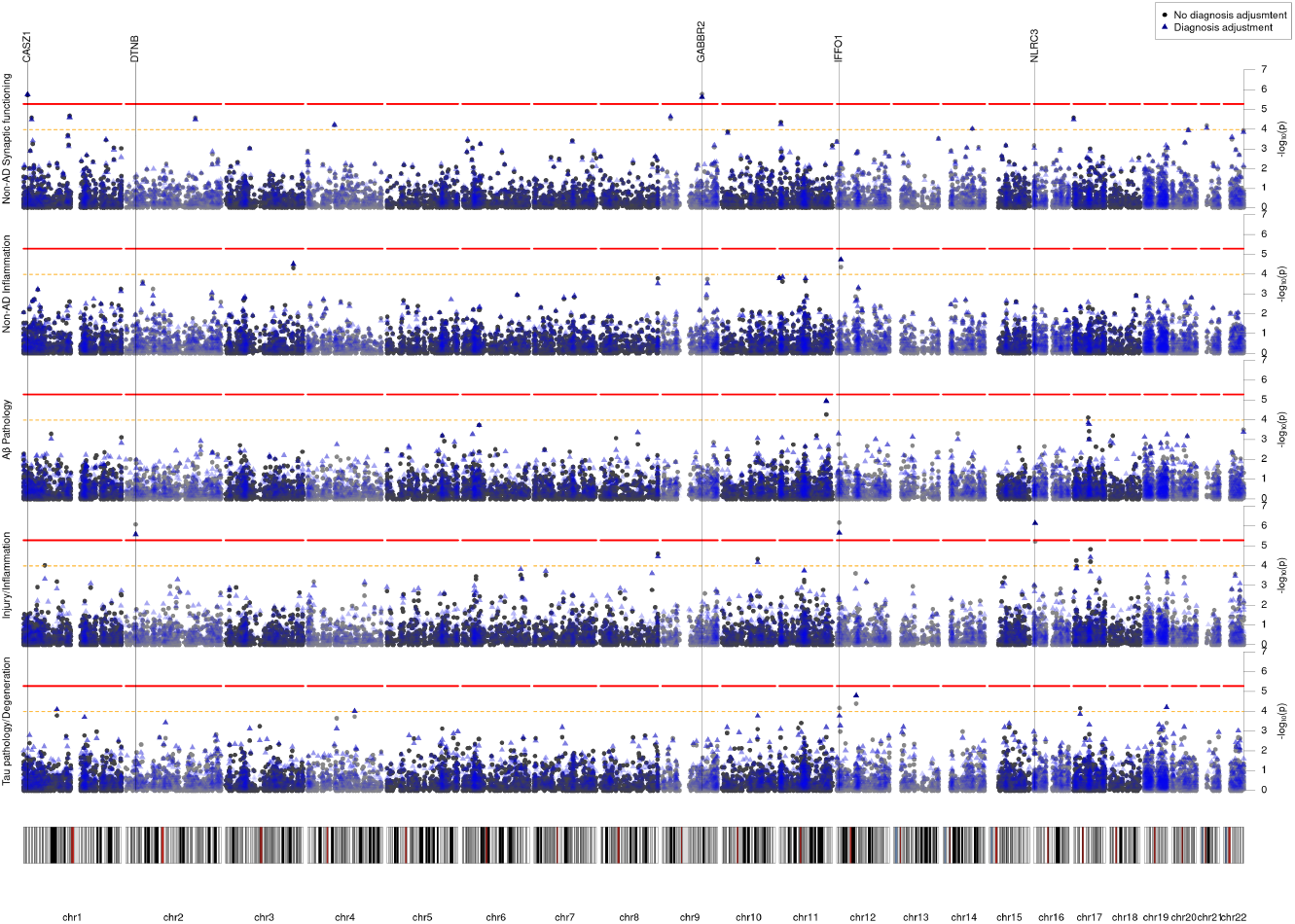
Manhattan plot of the exome-wide rare variant anayses (protein-coding). Results from the exome-wide rare variant (MAF < 1%) analyses of five CSF biomarker principal components (PC) (n=480). Each plot displays a different PC as outcome. X-axis represents each gene (rare protein-coding variants) and the y-axis the p-value obtained from gene-based SKAT-O tests on a -log_10_ scale. All analyses were adjusted for sex, age and genetic ancestry. Blue points represent p-values additionally adjusted for diagnosis. Red line indicates exome-wide significance threshold (p=5.2*10^−6^). Yellow line indicates suggestive threshold (p=1.0*10^−4^)

**Figure 2:**
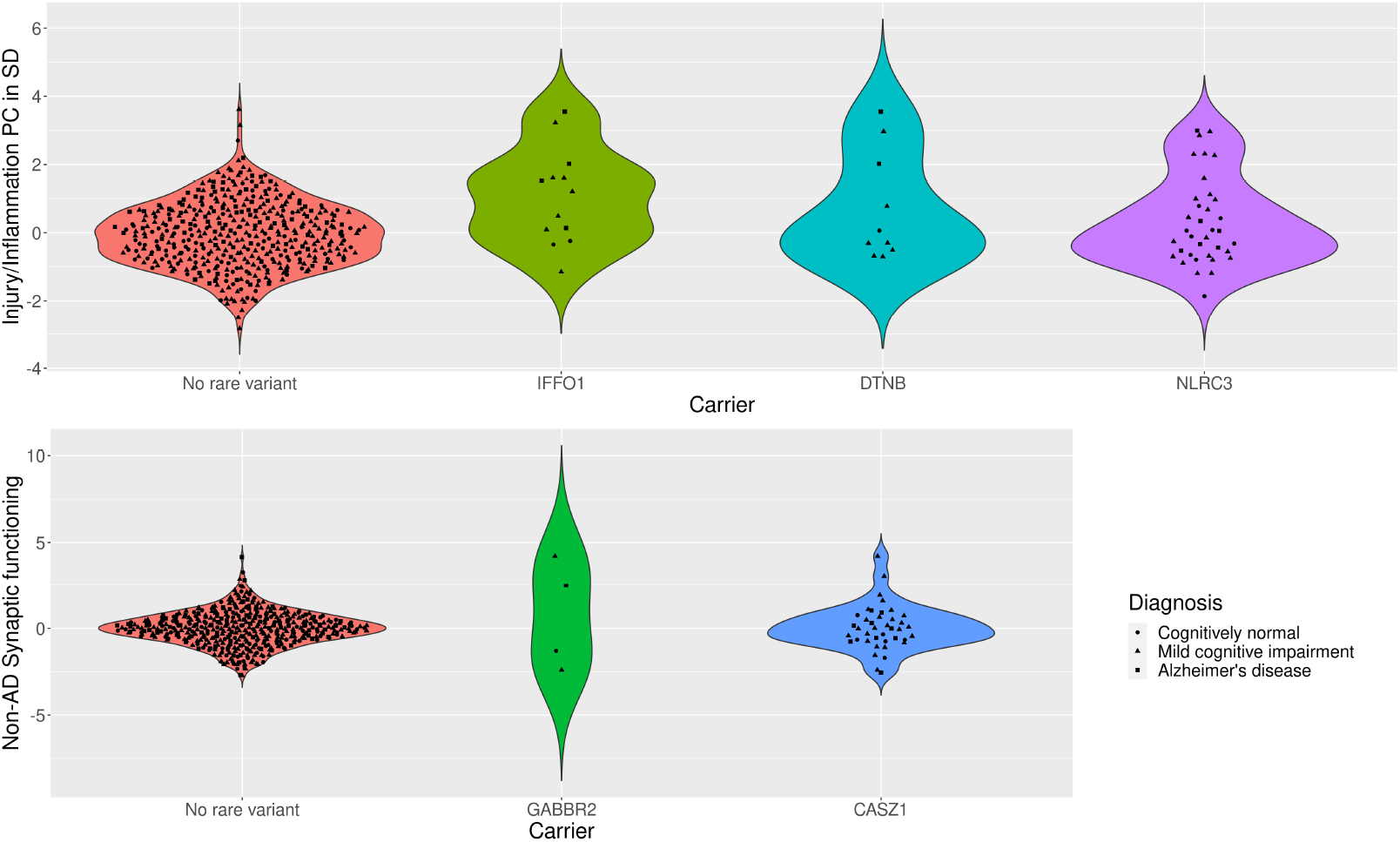
Violin plot of CSF biomarker principal component score distributions per rare variant carrier status. Top row displays the distribution of the Injury/Inflammation PC in participants not carrying a rare variant in the exome-wide significant genes, or carrying at least one variant in IFFO1, DTNB, or NLRC3. Bottom row displays the distribution of the Non-AD Synaptic functioning PC in participants not carrying a rare variant in the exome-wide significant genes, or carrying at least one variant in GABBR2, CASZ, or MICALCL. For the latter, only loss-of-function variants are considered, otherwise any protein-coding variant.

Two genes were associated at exome-wide significance with the Injury/Inflammation PC: *IFFO1* (p=6.7*10^−7^) and *DTNB* (p=8.3*10^−7^). *IFFO1* harbored five rare variants, results mostly being driven by rs138380449 and rs139792267, with five rare variant carriers each and a total of 10 carriers. The minor A allele in both SNPs was associated with 1.7SD (SE=0.44, p=.0001) and 1.3SD (SE=0.45, p=.0037) higher levels of injury/inflammation. Both SNPs are located six bp from each other in the *IFFO1* exon. The minor alleles are missense variants resulting in a proline-to-leucine substitution, predicted to be moderately deleterious (CADD>22.5). All but one carrier had either MCI (n=6) or AD (n=3). Mediation tests indicated that injury/inflammation levels affected by *IFFO1* variants would also affect dementia (p=9.5*10^−6^). For *DTNB*, rare variants were also associated with higher levels of injury/inflammation (Supplementary Table 2, Figure 2). Mediation tests were significant as well (p=0.001) and lookup of the total effect on MMSE revealed a nominally significant association (p=0.04). (Supplementary Table 3)

Two genes also associated with the non-AD synaptic functioning PC at exome-wide significance: *GABBR2* (p=1.6*10^−6^) and *CASZ1* (p=1.9*10^−6^). In contrast to the Injury/Inflammation associated genes, rare variants in these genes tended to be associated with both higher and lower levels (Supplementary Table 2, Figure 2). Given the low correlation between the non-AD synaptic functioning PC with MMSE, mediation tests were not significant (p=>0.82).

#### Diagnosis adjustment

When adjusting for diagnosis, one additional gene reached exome-wide significance: *NLRC3* as predictor of the Injury/Inflammation PC (p = 7.0*10^−7^). As with *IFFO1* and *DTNB*, rare variants on average showed higher PC scores, e.g., the T allele in rs61732418 was associated with 0.8SD (SE=0.41, p=0.04) higher levels based on six carriers, but is likely benign (CADD=0.1). As with the other Injury/Inflammation associated genes, the results suggest a mediation effect on dementia symptoms (p=0.002). See Supplementary Results for sensitivity analyses and single-cohort results.

### Whole-exome rare variant analysis: Loss-of-function variants

When restricting analyses to LoF variants, 270 genes remained, which passed QC and for which at least four participants had rare variants. The exome-wide significance threshold was therefore set to p<1.9*10^−4^. Most lambdas were 1.01 or lower (Supplementary Figure 1), except for Aβ Pathology, which was 1.1 and may indicate slight inflation. No gene passed exome-wide significance in LoF prioritized models (Supplementary Figure 2).

## Discussion

We performed the first multivariate exome-wide rare variant analysis of multiple AD CSF biomarkers in two multi-center studies. We observed a highly consistent clustering of the examined biomarkers into five independent components in both studies. We interpret the first component to represent tau pathology and neurodegeneration more generally, the second to indicate neuronal injury and inflammation, and the third component to represent Aβ pathology specifically. Not only did Aβ almost exclusively load on the third component, but Aβ also did not load on any other component. This suggests that Aβ represents a different disease process than the other biomarkers. E.g., Aβ accumulation is thought to precede first symptoms, whereas the other biomarkers are more representative of concurrent disease state.[9] The first three components correlated with dementia symptoms in the expected directions in both studies, however, the magnitude tended to be smaller in the ADNI study. The ADNI sample included only participants with MCI, resulting in higher mean MMSE scores and lower variability in the lower range, which may not generalize to clinical populations.

The fourth and fifth component loaded on YKL-40 and neurogranin, thus the first intuition may be to interpret these components as representing inflammation and synaptic functioning. However, these components did not correlate with dementia symptoms, which is at odds with previous analyses of these molecules. It is important to consider, that neurogranin and YKL-40 also loaded on the Tau/Neurodegeneration PC, and YKL-40 loaded also on the Injury/Inflammation PC. The fourth and fifth components may represent variation in inflammation and synaptic functioning, which is not related to dementia, the clinical variance being included in the first two components.

We then tested the contribution of rare variants towards the different disease processes identified by different combinations of biomarkers. *IFFO1, DTNB* and *NLRC3* were associated with the Injury/Inflammation PC, as represented by heightened Nfl and YKL-40 levels in the presence of rare variants in these genes. Notably, mediation tests suggested that these genes also affect dementia symptoms by impacting neural injury and inflammation. *IFFO1* codes for Intermediate filament family orphan 1 and is involved in DNA repair.[38] It is plausible that rare variants in *IFFO1* would affect Nfl levels, which are themselves an intermediate filament. The results suggest that rare variants in *IFFO1* affect sensitivity of the neuronal cytoskeleton to damage, resulting in neurodegeneration and potentially dementia symptoms.

*DTNB* encodes part of the dystrophin-associated protein complex (DAPC). DAPC links actin with extracellular space, is involved in cell signaling and has been mostly studied in the context of muscle diseases.[39] Post-analysis we became aware of another simultaneously conducted study by Prokopenko et al., who performed a region-based whole genome-sequencing association analysis on AD status in an independent dataset (NIMH/NIA ADSP).[40] Interestingly, rare variants in *DTNB* were associated with AD. The converging evidence from two independent studies, using a biomarker/pathway-based approach on the one hand, and a case-control design on the other, strongly suggests an involvement of *DTNB* in neurodegenerative processes and development of AD, not previously considered.

Finally, *NLRC3* was also associated with injury/inflammation, but only when statistically adjusting for diagnosis. However, the difference between both models was minor. *NLRC3* has a well-established role in lowering inflammation via inhibition of NfκB and NLRP3 inflammasome pathways, which have been observed to play a role in AD in human and mouse studies.[41] Specifically, a recent preprint reported that downregulation of *NLRC3* in a mouse model affects plaque deposition and neuronal loss.[42] Thus, we speculate that rare variants in *NLRC3* elevate inflammation, resulting in increased neurodegeneration and dementia symptoms.

*GABBR2* and *CASZ1* were genes identified with Non-AD synaptic functioning in protein-coding models, as mainly represented by higher levels of neurogranin. *GABBR2* encodes a GABA receptor, the main inhibitory neurotransmitter in the human brain. Downregulation of GABA receptors in various brain regions is associated with Alzheimer’s disease, potentially by disrupting the balance between excitation/inhibition balance.[43, 44] It seems plausible, that rare variants in the gene would also affect neurogranin levels and other markers of synaptic functioning. Curiously, despite prior evidence for an involvement of GABA in AD, we did not observe an association with dementia symptoms. Rare variants in the *GABBR2* gene therefore might only affect non-clinical variation of synaptic functioning, without consequences on neurodegeneration or dementia symptoms.

Finally, *CASZ1* is a zinc finger transcription factor expressed in the brain, but has been mostly studied in the context of cardiac health. For instance, LoF variants in the genes are associated with congenital heart disease[45] and cardiomyopathy[46].

Two genes reached exome-wide significance in the EMIF cohort only, but not in the mega-analysis: *CHI3L* and *CLU. CHI3L* encodes the YKL-40 protein, which is the primary biomarker loading on the non-AD inflammation PC and was recently identified as a cis-pQTL in a common-variant GWAS in an overlapping set of EMIF-AD MBD and ADNI individuals.[16] In regard to *CLU*, common and rare variants have been associated with AD and the gene product clusterin has been researched extensively as potential AD biomarker.[47, 48] Our results hint at *CLU* acting mostly via disruption of synaptic functioning, but the results have to be interpreted cautiously in light of non-replication.

The two biggest strengths of the study are the mega-analysis and multivariate design. The simultaneous analysis of multiple American and European centers and studies improves the generalizability of the results and allowed us to increase statistical power. The examination of biomarker combinations instead of single values likely supported the accurate and robust assessment of underlying disease processes, while improving power. This study is also one of the first to formally test mediation in the context of rare variant analyses using the recently developed SMUT approach.

However, as any other rare variant analysis, the chance for false positives or non-generalizable results is higher than for common variants. We opted for a mega-analysis instead of discovery-replication design to maximize robustness of initial findings. This choice also means that findings need to be externally verified before firm conclusions can be drawn. Another limitation of the study is that both CSF biomarkers and MMSE scores were measured concurrently and analyzed cross-sectionally in the mediation analyses. We can therefore not rule out reverse causation or independent pleiotropic gene effects on the biomarkers and dementia symptoms. A longitudinal analysis is recommended to explore gene effects further.

In summary, the results suggest that rare variants in *IFFO1, DTNB* and *NLRC3* impact neuronal injury and inflammation, by potentially altering cytoskeleton structure, impairing repair abilities and by disinhibition of immune pathways. The resulting sensitivity to damage and inflammation may then result in neurodegeneration and dementia symptoms. Finally, we also found evidence for the involvement of *GABBR2 and CASZ1* in synaptic functioning, but no evidence that these changes would impact dementia symptoms.

Supplementary information is available at MP’s website

## Data Availability

To comply with EU law and participant privacy, individual-level clinical data cannot be shared publicly, but can be requested via EMIF-AD.

https://emif-catalogue.eu

http://www.emif.eu/about/emif-ad

## EMIF

The authors thank the participants and families who took part in this research. The authors would also like to thank all people involved in data and sample collection and/or logistics across the different centres.

Funding: The present study was conducted as part of the EMIF-AD project, which has received support from the Innovative Medicines Initiative Joint Undertaking under EMIF grant agreement no. 115372, EPAD grant no. 115736, and from the European Union’s Horizon 2020 research and innovation programme under grant agreement No. 666992. Resources of which are composed of financial contribution from the European Union’s Seventh Framework Program (FP7/2007-2013) and EFPIA companies’ in-kind contribution. Research at VIB-UAntwerp was in part supported by the Research Foundation Flanders (FWO), and the University of Antwerp Research Fund, Belgium. FK is a supported by a PhD fellowship from the University of Antwerp Research Fund. The DESCRIPA study was funded by the European Commission within the fifth framework program (QLRT-2001-2455). The EDAR study was funded by the European Commission within the fifth framework program (contract no. 37670). The San Sebastian GAP study is partially funded by the Department of Health of the Basque Government (allocation 17.0.1.08.12.0000.2.454.01.41142.001.H). The Leuven cohort was funded by the Stichting voor Alzheimer Onderzoek (grant numbers #11020, #13007 and #15005). The Lausanne cohort was supported by grants from the Swiss National Research Foundation (SNF 320030_141179), Synapsis Foundation-Alzheimer Research Switzerland (grant number 2017-PI01).

HZ is a Wallenberg Scholar supported by grants from the Swedish Research Council (#2018-02532), the European Research Council (#681712), Swedish State Support for Clinical Research (#ALFGBG-720931), the Alzheimer Drug Discovery Foundation (ADDF), USA (#201809-2016862), the AD Strategic Fund and the Alzheimer’s Association (#ADSF-21-831376-C, #ADSF-21-831381-C and #ADSF-21-831377-C), the Olav Thon Foundation, the Erling-Persson Family Foundation, Stiftelsen för Gamla Tjänarinnor, Hjärnfonden, Sweden (#FO2019-0228), the European Union’s Horizon 2020 research and innovation programme under the Marie Skłodowska-Curie grant agreement No 860197 (MIRIADE), and the UK Dementia Research Institute at UCL.

FB is supported by the NIHR biomedical research centre at UCLH

## ADNI

Data collection and sharing for this project was funded by the Alzheimer’s Disease Neuroimaging Initiative (ADNI) (National Institutes of Health Grant U01 AG024904) and DOD ADNI (Department of Defense award number W81XWH-12-2-0012). ADNI is funded by the National Institute on Aging, the National Institute of Biomedical Imaging and Bioengineering, and through generous contributions from the following: AbbVie, Alzheimer’s Association; Alzheimer’s Drug Discovery Foundation; Araclon Biotech; BioClinica, Inc.; Biogen; Bristol-Myers Squibb Company; CereSpir, Inc.;Cogstate;Eisai Inc.; Elan Pharmaceuticals, Inc.; Eli Lilly and Company; EuroImmun; F. Hoffmann-La Roche Ltd and its affiliated company Genentech, Inc.; Fujirebio; GE Healthcare; IXICO Ltd.; Janssen Alzheimer Immunotherapy Research & Development, LLC.; Johnson & Johnson Pharmaceutical Research & Development LLC.; Lumosity; Lundbeck; Merck & Co., Inc.; Meso Scale Diagnostics, LLC.;NeuroRx Research; Neurotrack Technologies;Novartis Pharmaceuticals Corporation; Pfizer Inc.; Piramal Imaging; Servier; Takeda Pharmaceutical Company; and Transition Therapeutics. The Canadian Institutes of Health Research is providing funds to support ADNI clinical sites in Canada. Private sector contributions are facilitated by the Foundation for the National Institutes of Health (www.fnih.org). The grantee organization is the Northern California Institute for Research and Education, and the study is coordinated by the Alzheimer’s Therapeutic Research Institute at the University of Southern California. ADNI data are disseminated by the Laboratory for Neuro Imaging at the University of Southern California.

