## Supplementary Material for "Rare variants in *IFFO1, DTNB* and *NLRC3* associate with Alzheimer’s disease CSF profile of neuronal injury and inflammation"

**AUTHORS**

Alexander Neumann PhD, Fahri Küçükali Msc, Isabelle Bos PhD, Stephanie J.B. Vos PhD, Sebastiaan Engelborghs PhD, Tim De Pooter BSc, Geert Joris BSc, Peter De Rijk PhD, Ellen De Roeck PhD, Magda Tsolaki MD, PhD, Frans Verhey PhD, Pablo Martinez-Lage MD, Mikel Tainta MD, Giovanni Frisoni PhD, Oliver Blin MD, PhD, Jill Richardson PhD, Régis Bordet PhD, Philip Scheltens MD, Julius Popp MD, Gwendoline Peyratout PhD, Peter Johannsen MD, PhD, Lutz Frölich MD, PhD, Rik Vandenberghe PhD, Yvonne Freund-Levi PhD, Johannes Streffer MD, PhD, Simon Lovestone PhD, Cristina Legido-Quigley PhD, Mara ten Kate PhD, Frederik Barkhof MD, PhD, Mojca Strazisar PhD, Henrik Zetterberg MD, PhD, Lars Bertram MD, Pieter Jelle Visser MD, PhD, Christine van Broeckhoven PhD, Kristel Sleegers MD, PhD

#### CORRESPONDING AUTHOR

Alexander Neumann

VIB Center for Molecular Neurology

Universiteitsplein 1

B-2610 Antwerp

Belgium

### Supplementary Methods

#### Participant Selection

In total 450 participants were genotyped in the EMIF study using exome-wide sequencing and 808 in the ADNI study using whole-exome sequencing. Participant selection was based on availability and quality of DNA samples, sample volume (100ng/µl in 25µl), no known inconsistencies, and secondary QC for WES library prep. We excluded participants with known pathogenic mutations: five individuals in EMIF had mutations in *PSEN1*, *APP* and *GRN*, and three individuals in ADNI had mutations in *PSEN1* and *MAPT*. Furthermore, we excluded one family member per related participant (defined as PI-HAT > 0.4 in IBD analysis), to avoid confounding by shared environment. Participants with more complete information were prioritized or, in case of equal data completeness, randomly selected. In EMIF three participants and in ADNI five participants were removed due to relatedness with another member in the sample.

For the main analysis we selected participants, who were assessed with exome-wide sequencing, had no known pathogenic mutations, were unrelated and had information on at most one CSF biomarker missing. The main analysis sample for EMIF was 353 and for ADNI 127, resulting in a total sample size of 480 participants. Two EMIF participants had missing mini-mental state examination (MMSE) information, leading to a slightly reduced sample size for the mediation analysis (n=478). For lookup of MMSE associations, which require only MMSE and no biomarker information, the sample size was larger with EMIF contributing 440 participants and ADNI

800, resulting in a total sample size of 1240 participants. Six participants had non-European ancestry and were excluded in sensitivity analyses to test the robustness of the results to population stratification.

#### Genotyping

Whole exome-sequencing in EMIF was performed by the Neuromics Support Facility (NSF) at the VIB-UAntwerp Center for Molecular Neurology, Antwerp, Belgium, on Illumina NextSeq500 platform using paired-end reads on DNA samples hybridized with SeqCap EZ Human Exome Kit v3.0 (Roche). Per run, a maximum of 12 samples were run, resulting in an average of 91 million reads per sample. On average 93.85% of targeted sites minimum 20X coverage per sample and on average 30-40X. In ADNI whole exome-sequencing was performed using the Illumina HiSeq2000 platform on DNA samples hybridized with the Agilent's SureSelect Human All Exon 50 Mb kit, see previous publications for details.[1] The same quality control pipeline was then applied to both studies.

After demultiplexing and trimming adaptors with fastq-mcf 1.1.2-537,[2] read quality was assessed with FastQC 0.11.5 and aligned to the hg19 human reference genome with the Burrows-Wheeler Aligner 0.7.15.[3] Samtools was used for duplicate removal.[4] Gender checks were performed with GenomeComb.[5] and variant calling with the Genome Analysis Toolkit 4.0.6.[6] Specifically we applied GATK HaplotypeCaller, GenomicsDBImport and GenotypeGVCFs to create a raw .vcf file,[7] then the Variant Quality Score Recalibration tool was used to filter SNPs not in the 99.5% (EMIF) or 99.7% (ADNI) and indels not in the 99% sensitivity tranche. Multi-allelic variants were represented in different rows, and indels were left-aligned and normalized using bcftools norm. Annotation was performed with bcftools using dbSNP Build 151. Genotypes with a genotype quality<20, depth<8, allele depth>1:3 (heterozygous variants) or 1:9 (homozgous variants) were excluded with the help of VCFtools 0.1.16 and vcffilterjdk.[8] Furthermore, variants with deviation of Hardy-Weinberg Equilibrium ($p<10^{-7}$), genotype quality<35 or call rate<80% were also removed.[9] Annotations were performed with SnpEff[10], ANNOVAR[11] and CADD 1.4[12]. None of the remaining samples had a call rate below 95%. X-chromosome heterozygosity rates were within expected ranges given the reported sex. Genetic ancestry and relatedness was assessed using high-quality LD-pruned common variants (PLINK argument "--indep-pairwise 50 5 0.5", MAF > 1%, call rate > 98%). Genetic ancestry was estimated using Somalier[13] and the super populations from the 1000 Genomes reference[14], with participants having less than 80% European ancestry proportion classified as having non-European ancestry. Furhtermore, principal components were estimated and used as covariates in the analysis.

Post-analysis, we retained only genes with at least two rare variant carriers in each study to reduce Type-1 error. Furthermore, we excluded the USP30 gene due to convergence problems resulting in extremely low p-values ($p<10^{-100}$) for all outcomes.

#### Statistical analysis

For single variant analyses, we used a multiple imputation approach with MICE (40 imputations, 20 iterations) to impute missing genotype data, as opposed to the default mean genotype imputation in MetaSKAT tests.[15]

### Supplementary Results

#### Sensitivity analyses and single-cohort results

When restricting the analysis sample to participants with European ancestry, results stayed mostly consistent (Supplementary Table S2). Two additional genes, CLCNKB and SLC22A10, were exome-wide significant in LoF prioritized models, when restricting to European ancestry. CLCNKB was significantly associated with non-AD inflammation ($p=2.2E^{-04}$), when not adjusting for diagnosis, SLC22A10 was significantly associated with injury/inflammation, but only when adjusting for diagnosis. Due to these inconsistent associations across models and samples, we do not discuss these findings further.

The primary outcome of this study was the mega-analysis results, as they are expected to be more robust. Supplementary Table S4 and S5 lists genes, which were exome-wide significant in one of the studies, but not in joint analyses. While these genes generally did not replicate, when taking into account multiple testing, we wish to highlight two exome-wide significant genes in EMIF: *CHI3L1* associated with non-AD Inflammation ($p_{EMIF}=4.9*10^{-8}$, $p_{ADNI}=0.07$,$p_{meta}=0.02$) and *CLU* with non-AD Synaptic functioning ($p_{EMIF}=6.0*10^{-7}$, $p_{ADNI}=0.45$,$p_{meta}=0.01$).

### Supplementary Tables

#### Table S1: PCA loadings in analysis sample vs full sample

| | Tau pathology/ Degeneration | | Injury/ Inflammation | | Aβ Pathology | | Non-AD Inflammation | | Non-AD Synaptic functioning | |
|---|---|---|---|---|---|---|---|---|---|---|
| Sample | Analysis (n=480) | Full (n=1158) | Analysis (n=480) | Full (n=1158) | Analysis (n=480) | Full (n=1158) | Analysis (n=480) | Full (n=1158) | Analysis (n=480) | Full (n=1158) |
| *Loadings* | | | | | | | | | | |
| Tau | 0.86 | 0.87 | 0.23 | 0.23 | -0.05 | -0.06 | 0.27 | 0.24 | 0.28 | 0.27 |
| pTau | 0.92 | 0.91 | 0.11 | 0.14 | -0.07 | -0.08 | 0.17 | 0.20 | 0.26 | 0.26 |
| Aβ | -0.07 | -0.07 | -0.04 | -0.03 | 1.00 | 1.00 | -0.04 | 0.00 | 0.03 | 0.04 |
| NFL | 0.19 | 0.21 | 0.94 | 0.94 | -0.05 | -0.03 | 0.25 | 0.25 | 0.09 | 0.08 |
| YKL-40 | 0.31 | 0.32 | 0.30 | 0.32 | -0.05 | 0.00 | 0.89 | 0.88 | 0.16 | 0.17 |
| Ng | 0.48 | 0.51 | 0.11 | 0.10 | 0.05 | 0.07 | 0.18 | 0.18 | 0.85 | 0.83 |

**Table 1: PCA results.** Principal component analysis of CSF biomarkers in main analysis sample with genetic information available (n=480) and in full sample not filtered for availability of genetic information (n=1158). Component loadings of each biomarker (first column) on five principal components (column groups two to six) are displayed.

#### Table S2: Single-variant results (spreadsheet download separately available).

This table contains the association results of single variants, as well as additional information on location, function and predicted deleteriousness. We only considered variants within genes showing exome-wide significance (Table 3) and which were polymorphic in the analysis sample (max n=480). Depending on whether the gene was significant in a protein-coding or loss-of-function model, we only analyzed rare (MAF <1%) protein-coding or loss-of-function variants, respectively. We associated these variants with those CSF biomarker principal components (PC), which showed a significant association on a gene level. PCs, residualized for sex, age and genetic ancestry, were regressed on the number of rare alleles.

#### Table S3: Associations between exome-wide significant genes and MMSE

| | | | ADNI | | | | EMIF | | | | Mega | | | | |
|---|---|---|---|---|---|---|---|---|---|---|---|---|---|---|---|
| Mediator | Gene | Model | n | $n_{snps}$ | $n_{carriers}$ | p | n | $n_{snps}$ | $n_{carriers}$ | p | n | $n_{snps}$ | $N_{snps\ overlap}$ | $n_{carriers}$ | p |
| Injury/ Inflammation | IFFO1 | protein | 800 | 6 | 30 | 1.00 | 440 | 4 | 13 | 0.14 | 1240 | 8 | 2 | 43 | 0.74 |
| | DTNB | protein | 800 | 20 | 27 | 0.50 | 440 | 4 | 7 | 0.05 | 1240 | 22 | 2 | 34 | 0.04 |
| | NLRC3 | protein | 800 | 43 | 72 | 0.75 | 440 | 23 | 33 | 0.32 | 1240 | 59 | 7 | 106 | 0.49 |
| Non-AD Synaptic functioning | GABBR2 | protein | 800 | 5 | 5 | 0.83 | 440 | 3 | 3 | 0.27 | 1240 | 8 | 0 | 8 | 0.51 |
| | CASZ1 | protein | 800 | 43 | 86 | 0.38 | 440 | 21 | 34 | 0.21 | 1240 | 57 | 7 | 120 | 0.53 |

**Table S2: Associations between exome-wide significant genes and MMSE.** This table shows whether evidence of association was found between MMSE and any of the genes identified to show exome-wide significant association with a biomarker PC.
**Model** Indicator whether variants were restricted to protein-coding (Protein) or loss-of-function (LoF) variants
**n** Sample size
**$n_{snps}$** Number of variants included

$\mathbf{n}_{\mathbf{carriers}}$ Number of participants with at least one rare variant in the gene
**p** p-value of SKAT-O test
$\mathbf{n}_{\mathbf{snp\ overlap}}$ Number of variants present in both ADNI and EMIF

### Table S4: Exome-wide significant genes in European ancestry subsample

| | | | ADNI | | | | EMIF | | | | Mega | | | | |
|---|---|---|---|---|---|---|---|---|---|---|---|---|---|---|---|
| Outcome | Gene | Model | n | $n_{snps}$ | $n_{carriers}$ | p | n | $n_{snps}$ | $n_{carriers}$ | p | n | $n_{snps}$ | $N_{snps}$ overlap | $n_{carriers}$ | p |
| *Adjusted for sex, age and ancestry* | | | | | | | | | | | | | | | |
| Tau pathology/Degeneration | - | - | - | - | - | - | - | - | - | - | - | - | - | - | - |
| Injury/Inflammation | IFFO1 | protein | 121 | 6 | 4 | 5.6E-01 | 353 | 4 | 9 | 2.0E-06 | 474 | 8 | 2 | 13 | 7.6E-07 |
| | DTNB | protein | 121 | 20 | 2 | 6.0E-01 | 353 | 4 | 7 | 9.1E-04 | 474 | 22 | 2 | 9 | 8.3E-07 |
| Aβ Pathology | - | - | - | - | - | - | - | - | - | - | - | - | - | - | - |
| Non-AD Inflammation | CLCNKB | LoF- | 121 | 3 | 3 | 4.8E-04 | 353 | 2 | 2 | 3.8E-02 | 474 | 5 | 0 | 5 | 2.2E-04 |
| Non-AD Synaptic functioning | CASZ1 | protein | 121 | 43 | 11 | 4.2E-01 | 353 | 2 | 2 | 2.2E-04 | 474 | 57 | 7 | 40 | 1.9E-06 |
| | GABBR2 | protein | 121 | 5 | 2 | 5.4E-01 | 353 | 3 | 2 | 4.5E-06 | 474 | 8 | 0 | 4 | 4.7E-06 |
| *Adjusted for sex, age, ancestry and diagnosis* | | | | | | | | | | | | | | | |
| Tau pathology/Degeneration | - | - | - | - | - | - | - | - | - | - | - | - | - | - | - |
| Injury/Inflammation | NLRC3 | protein | 121 | 43 | 9 | 2.2E-01 | 353 | 24 | 25 | 1.2E-05 | 474 | 60 | 7 | 34 | 1.9E-06 |
| | IFFO1 | protein | 121 | 6 | 4 | 7.4E-01 | 353 | 4 | 9 | 9.2E-06 | 474 | 8 | 2 | 13 | 2.5E-06 |
| | DTNB | protein | 121 | 20 | 2 | 4.5E-01 | 353 | 4 | 7 | 1.4E-03 | 474 | 22 | 2 | 9 | 2.6E-06 |
| | SLC22A10 | LoF | 121 | 4 | 2 | 1 | 353 | 2 | 3 | 1.2E-04 | 474 | 5 | 1 | 5 | 2.1E-04 |
| Aβ Pathology | - | - | - | - | - | - | - | - | - | - | - | - | - | - | - |
| Non-AD Inflammation | - | - | - | - | - | - | - | - | - | - | - | - | - | - | - |
| Non-AD Synaptic functioning | CASZ1 | protein | 121 | 43 | 11 | 4.5E-01 | 353 | 21 | 29 | 9.5E-06 | 474 | 57 | 7 | 40 | 1.6E-06 |

**Table S3: Exome-wide significant genes in European ancestry subsample.** Results for exome-wide rare-variant and mediation analyses, when analysis sample is restricted to participants of European ancestry. Rare (MAF<1%) protein-coding variants in 13,363 genes were tested on a gene level in the protein-coding model and 536 genes in the loss-of-function model. Each gene was associated with five principal component scores of CSF biomarkers, representing different neurodegenerative processes.  P-values (p) were obtained from gene-based SKAT-O tests. SMUT tested mediation on dementia symptoms (MMSE scores) via changes in the principal components All tests were adjusted for sex, age and genetic ancestry (top group). In separate models, we additionally adjusted for diagnosis status (bottom group). Only genes with exome-wide significant association in the mega-analysis (Mega) are displayed (protein-coding:  $p<5.2*10^{-6}$; lof:  $p<1.9*10^{-4)}$)
**Model** Indicator whether variants were restricted to protein-coding (Protein) or loss-of-function (LoF) variants
**n** Sample size
$\mathbf{n_{snps}}$ Number of variants included
$\mathbf{n_{carriers}}$ Number of participants with at least one rare variant in the gene
**p** p-value of SKAT-O test.
$\mathbf{n_{snp\ overlap}}$ Number of variants present in both ADNI and EMIF

### Table S5: Genes with exome-wide significance in single cohort (Protein-coding)

| | | ADNI | | | | EMIF | | | | Meta | | | | |
|---|---|---|---|---|---|---|---|---|---|---|---|---|---|---|
| Outcome | Gene | n | $n_{snps}$ | $n_{carriers}$ | p | n | $n_{snps}$ | $n_{carriers}$ | p | n | $n_{snps}$ | $N_{snps\ overlap}$ | $n_{carriers}$ | p |
| *Adjusted for sex, age and ancestry* | | | | | | | | | | | | | | |
| Tau pathology/Degeneration | NCAM2 | 127 | 11 | 2 | 3.1E-06 | 353 | 4 | 3 | 1.7E-01 | 480 | 15 | 0 | 5 | 3.6E-03 |
| Injury/Inflammation | - | - | - | - | - | - | - | - | - | - | - | - | - | - |
| Aβ Pathology | - | - | - | - | - | - | - | - | - | - | - | - | - | - |
| Non-AD Inflammation | PRPS1L1 | 127 | 7 | 2 | 3.5E-06 | 353 | 3 | 2 | 7.9E-01 | 480 | 8 | 2 | 4 | 4.8E-03 |
| | CHI3L1 | 127 | 16 | 4 | 1.1E-01 | 353 | 8 | 10 | 1.2E-07 | 480 | 20 | 4 | 14 | 1.5E-02 |
| Non-AD Synaptic functioning | CLU | 127 | 7 | 1 | 4.6E-01 | 353 | 6 | 4 | 5.9E-07 | 480 | 10 | 3 | 5 | 1.0E-02 |
| *Adjusted for sex, age, ancestry and diagnosis* | | | | | | | | | | | | | | |
| *Adjusted for sex, age and ancestry* | | | | | | | | | | | | | | |
| Tau pathology/Degeneration | AMBN | 127 | 13 | 3 | 1.7E-30 | 353 | 7 | 6 | 7.7E-01 | 480 | 18 | 2 | 9 | 1 |
| | COQ6 | 127 | 14 | 2 | 2.6E-30 | 353 | 7 | 7 | 9.1E-02 | 480 | 18 | 3 | 9 | 6.4E-02 |
| | MAST4 | 127 | 40 | 8 | 1.9E-06 | 353 | 31 | 33 | 2.6E-01 | 480 | 67 | 4 | 43 | 5.9E-02 |
| | POGLUT1 | 127 | 9 | 2 | 4.0E-28 | 353 | 2 | 4 | 4.3E-01 | 480 | 10 | 1 | 6 | 9.3E-02 |
| | SRF | 127 | 7 | 2 | 6.3E-29 | 353 | 4 | 5 | 1 | 480 | 9 | 2 | 7 | 8.3E-01 |
| | TMEM261 | 127 | 4 | 3 | 2.4E-28 | 353 | 4 | 3 | 1.8E-01 | 480 | 7 | 1 | 6 | 7.4E-01 |
| | ZFYVE21 | 127 | 4 | 2 | 4.0E-28 | 353 | 2 | 2 | 2.5E-01 | 480 | 5 | 1 | 4 | 5.3E-02 |
| Injury/Inflammation | AMBN | 127 | 13 | 3 | 3.9E-11 | 353 | 7 | 6 | 1 | 480 | 18 | 2 | 9 | 5.7E-01 |
| | COQ6 | 127 | 14 | 2 | 4.8E-11 | 353 | 7 | 7 | 1.0E-02 | 480 | 18 | 3 | 9 | 3.9E-02 |
| | POGLUT1 | 127 | 9 | 2 | 4.8E-11 | 353 | 2 | 4 | 1.8E-01 | 480 | 10 | 1 | 6 | 1.5E-01 |

| | | | | | | | | | | | | | | |
|---|---|---|---|---|---|---|---|---|---|---|---|---|---|---|
| | SRF | 127 | 7 | 2 | 3.6E-11 | 353 | 4 | 5 | 6.3E-01 | 480 | 9 | 2 | 7 | 8.2E-01 |
| | TMEM261 | 127 | 4 | 3 | 5.0E-11 | 353 | 4 | 3 | 8.6E-01 | 480 | 7 | 1 | 6 | 1 |
| | TXNRD2 | 127 | 13 | 3 | 4.2E-11 | 353 | 1 | 1 | 5.0E-01 | 480 | 13 | 1 | 4 | 5.0E-01 |
| | ZFYVE21 | 127 | 4 | 2 | 3.9E-11 | 353 | 2 | 2 | 8.1E-01 | 480 | 5 | 1 | 4 | 2.8E-01 |
| Aβ Pathology | APOBEC3B | 127 | 9 | 3 | 1.9E-11 | 353 | 4 | 5 | 2.9E-01 | 480 | 12 | 1 | 8 | 2.5E-01 |
| | COLCA1 | 127 | 4 | 2 | 1.3E-07 | 353 | 1 | 1 | 4.9E-01 | 480 | 4 | 1 | 3 | 5.2E-01 |
| | GPAM | 127 | 10 | 2 | 1.3E-08 | 353 | 6 | 3 | 2.1E-01 | 480 | 16 | 0 | 5 | 2.6E-01 |
| | MMP7 | 127 | 4 | 2 | 2.2E-10 | 353 | 2 | 2 | 5.6E-01 | 480 | 5 | 1 | 4 | 5.9E-01 |
| | SERPINA11 | 127 | 6 | 2 | 6.4E-09 | 353 | 5 | 6 | 8.2E-01 | 480 | 9 | 2 | 8 | 3.3E-01 |
| | UTS2R | 127 | 10 | 2 | 1.3E-07 | 353 | 2 | 2 | 5.0E-02 | 480 | 12 | 0 | 4 | 4.9E-02 |
| Non-AD Inflammation | AMBN | 127 | 13 | 3 | 7.0E-13 | 353 | 7 | 6 | 3.7E-01 | 480 | 18 | 2 | 9 | 2.1E-01 |
| | APOBEC3B | 127 | 9 | 3 | 5.1E-21 | 353 | 4 | 5 | 4.3E-01 | 480 | 12 | 1 | 8 | 8.4E-02 |
| | COLCA1 | 127 | 4 | 2 | 7.4E-24 | 353 | 1 | 1 | 6.4E-01 | 480 | 4 | 1 | 3 | 6.9E-01 |
| | COQ6 | 127 | 14 | 2 | 7.4E-13 | 353 | 7 | 7 | 4.8E-01 | 480 | 18 | 3 | 9 | 5.9E-01 |
| | GPAM | 127 | 10 | 2 | 5.2E-22 | 353 | 6 | 3 | 1 | 480 | 16 | 0 | 5 | 7.6E-01 |
| | MMP7 | 127 | 4 | 2 | 7.2E-32 | 353 | 2 | 2 | 6.7E-01 | 480 | 5 | 1 | 4 | 1 |
| | POGLUT1 | 127 | 9 | 2 | 7.7E-13 | 353 | 2 | 4 | 9.7E-01 | 480 | 10 | 1 | 6 | 5.4E-01 |
| | RALGPS2 | 127 | 4 | 2 | 2.7E-06 | 353 | 2 | 4 | 2.0E-01 | 480 | 5 | 1 | 6 | 1.8E-03 |
| | SERPINA11 | 127 | 6 | 2 | 7.2E-23 | 353 | 5 | 6 | 2.9E-01 | 480 | 9 | 2 | 8 | 6.7E-02 |
| | SRF | 127 | 7 | 2 | 1.7E-12 | 353 | 4 | 5 | 2.0E-01 | 480 | 9 | 2 | 7 | 8.3E-01 |
| | TMEM261 | 127 | 4 | 3 | 5.6E-13 | 353 | 4 | 3 | 7.3E-01 | 480 | 7 | 1 | 6 | 2.6E-01 |
| | UTS2R | 127 | 10 | 2 | 1.2E-24 | 353 | 2 | 2 | 4.3E-01 | 480 | 12 | 0 | 4 | 5.8E-01 |
| | TXNRD2 | 127 | 13 | 3 | 7.8E-13 | 353 | 1 | 1 | 1.9E-01 | 480 | 13 | 1 | 4 | 1.9E-01 |
| | ZFYVE21 | 127 | 4 | 2 | 1.0E-12 | 353 | 2 | 2 | 5.7E-01 | 480 | 5 | 1 | 4 | 4.4E-02 |
| | CHI3L1 | 127 | 16 | 4 | 7.2E-02 | 353 | 8 | 10 | 4.9E-08 | 480 | 20 | 4 | 14 | 1.7E-02 |

| | | | | | | | | | | | | | | |
|---|---|---|---|---|---|---|---|---|---|---|---|---|---|---|
| Non-AD Synaptic functioning | AMBN | 127 | 13 | 3 | 3.1E-08 | 353 | 7 | 6 | 8.5E-01 | 480 | 18 | 2 | 9 | 4.8E-01 |
| | APOBEC3B | 127 | 9 | 3 | 2.3E-44 | 353 | 4 | 5 | 7.8E-01 | 480 | 12 | 1 | 8 | 1 |
| | COLCA1 | 127 | 4 | 2 | 2.7E-27 | 353 | 1 | 1 | 2.3E-01 | 480 | 4 | 1 | 3 | 2.4E-01 |
| | COQ6 | 127 | 14 | 2 | 9.6E-10 | 353 | 7 | 7 | 4.5E-02 | 480 | 18 | 3 | 9 | 4.2E-02 |
| | GPAM | 127 | 10 | 2 | 4.0E-35 | 353 | 6 | 3 | 7.4E-01 | 480 | 16 | 0 | 5 | 8.6E-01 |
| | MMP7 | 127 | 4 | 2 | 3.0E-46 | 353 | 2 | 2 | 1 | 480 | 5 | 1 | 4 | 6.4E-01 |
| | POGLUT1 | 127 | 9 | 2 | 5.8E-10 | 353 | 2 | 4 | 3.6E-01 | 480 | 10 | 1 | 6 | 1.7E-01 |
| | SERPINA11 | 127 | 6 | 2 | 9.7E-42 | 353 | 5 | 6 | 2.4E-01 | 480 | 9 | 2 | 8 | 9.2E-02 |
| | SRF | 127 | 7 | 2 | 6.4E-08 | 353 | 4 | 5 | 7.0E-01 | 480 | 9 | 2 | 7 | 5.5E-01 |
| | TMEM261 | 127 | 4 | 3 | 6.2E-08 | 353 | 4 | 3 | 9.4E-01 | 480 | 7 | 1 | 6 | 1 |
| | TXNRD2 | 127 | 13 | 3 | 6.5E-09 | 353 | 1 | 1 | 1.9E-01 | 480 | 13 | 1 | 4 | 1.9E-01 |
| | UTS2R | 127 | 10 | 2 | 6.7E-28 | 353 | 2 | 2 | 7.2E-01 | 480 | 12 | 0 | 4 | 6.8E-01 |
| | ZFYVE21 | 127 | 4 | 2 | 1.4E-08 | 353 | 2 | 2 | 1 | 480 | 5 | 1 | 4 | 3.6E-01 |
| | CLU | 127 | 7 | 1 | 4.5E-01 | 353 | 6 | 4 | 6.0E-07 | 480 | 10 | 3 | 5 | 1.2E-02 |

**Table S4: Genes with exome-wide significance in single cohort (Protein-coding).** Results for exome-wide rare-variant and mediation analyses. Rare (MAF<1%) protein-coding variants in 13,363 genes were tested on a gene level in the protein-coding model. Each gene was associated with five principal component scores of CSF biomarkers, representing different neurodegenerative processes. P-values (p) were obtained from gene-based SKAT-O tests. SMUT tested mediation on dementia symptoms (MMSE scores) via changes in the principal components. All tests were adjusted for sex, age and genetic ancestry (top group). In separate models, we additionally adjusted for diagnosis status (bottom group). Only genes with exome-wide significant association in one of the two studies, but not mega-analysis, are displayed (protein-coding: $p<5.2*10^{-6}$; lof: $p<1.9*10^{-4}$).

**n** Sample size

$\mathbf{n}_{snps}$ Number of variants included

$\mathbf{n}_{carriers}$ Number of participants with at least one rare variant in the gene

**p** p-value of SKAT-O test.

$\mathbf{n}_{snp\ overlap}$ Number of variants present in both ADNI and EMIF

### Table S6: Genes with exome-wide significance in single cohort (Loss-of-function)

| | | ADNI | | | | EMIF | | | | Meta | | | | |
|---|---|---|---|---|---|---|---|---|---|---|---|---|---|---|
| Outcome | Gene | n | $n_{snps}$ | $n_{carriers}$ | p | n | $n_{snps}$ | $n_{carriers}$ | p | n | $n_{snps}$ | $N_{snps\ overlap}$ | $n_{carriers}$ | p |
| *Adjusted for sex, age and ancestry* | | | | | | | | | | | | | | |
| Tau pathology/Degeneration | SP100 | 127 | 3 | 2 | 8.1E-87 | 353 | 2 | 3 | 2.9E-01 | 480 | 3 | 1 | 5 | 3.0E-01 |
| Injury/Inflammation | - | - | - | - | - | - | - | - | - | - | - | - | - | - |
| Aβ Pathology | - | - | - | - | - | - | - | - | - | - | - | - | - | - |
| Non-AD Inflammation | SP100 | 127 | 3 | 2 | 3.2E-09 | 353 | 1 | 3 | 1.1E-01 | 480 | 3 | 1 | 5 | 9.7E-02 |
| Non-AD Synaptic functioning | SP100 | 127 | 3 | 2 | 1.3E-18 | 353 | 1 | 3 | 4.4E-01 | 480 | 3 | 1 | 5 | 4.6E-01 |
| *Adjusted for sex, age, ancestry and diagnosis* | | | | | | | | | | | | | | |
| Tau pathology/Degeneration | SP100 | 127 | 3 | 2 | 1.9E-06 | 353 | 1 | 3 | 5.8E-01 | 480 | 3 | 1 | 5 | 6.1E-01 |
| Injury/Inflammation | DCHS2 | 127 | 6 | 2 | 2.8E-11 | 353 | 4 | 8 | 1 | 480 | 8 | 2 | 10 | 7.6E-01 |
| Aβ Pathology | SP100 | 127 | 3 | 2 | 2.4E-79 | 353 | 1 | 3 | 5.8E-01 | 480 | 3 | 1 | 5 | 6.1E-01 |
| Non-AD Inflammation | SP100 | 127 | 3 | 2 | 2.1E-10 | 353 | 1 | 3 | 6.9E-02 | 480 | 3 | 1 | 5 | 6.2E-02 |
| Non-AD Synaptic functioning | DCHS2 | 127 | 6 | 2 | 7.8E-26 | 353 | 4 | 8 | 3.7E-01 | 480 | 8 | 2 | 10 | 2.9E-01 |
| | SP100 | 127 | 3 | 2 | 4.3E-18 | 353 | 1 | 3 | 3.1E-01 | 480 | 3 | 1 | 5 | 3.2E-01 |

**Table S5: Genes with exome-wide significance in single cohort (Loss-of-function).** Results for exome-wide rare-variant and mediation analyses. Rare (MAF<1%) loss-of-function variants in 536 genes were tested on a gene level in the protein-coding model. Each gene was associated with five principal component scores of CSF biomarkers, representing different neurodegenerative processes. P-values (p) were obtained from gene-based SKAT-O tests. SMUT tested mediation on dementia symptoms (MMSE scores) via changes in the principal components. All tests were adjusted for sex, age and genetic ancestry (top group). In separate models, we additionally adjusted for diagnosis status (bottom group). Only genes with exome-wide significant association in one of the two studies, but not mega-analysis, are displayed (protein-coding: $p<5.2*10^{-6}$; lof: $p<1.9*10^{-4}$)).

**n** Sample size

**$n_{snps}$** Number of variants included

**$n_{carriers}$** Number of participants with at least one rare variant in the gene

**p** p-value of SKAT-O test.
$\mathbf{n}_{\textbf{snp overlap}}$ Number of variants present in both ADNI and EMIF

### Supplementary Figures

**Figure S1: QQ-plot of exome-wide rare variant analyses.** Colored points represent p-value distribution of the exome-wide rare variant (MAF < 1%) analyses of the five CSF biomarker principal components obtained from gene-based SKAT-O tests (n=480). The diagonal line together with its 95% confidence interval (grey area) represents the expected p-value distribution assuming chance finding. First row represents results from protein-coding variants. The bottom row represents results from loss-of-function variants. All analyses were adjusted for sex, age and genetic ancestry. The right column was additionally adjusted for diagnosis.

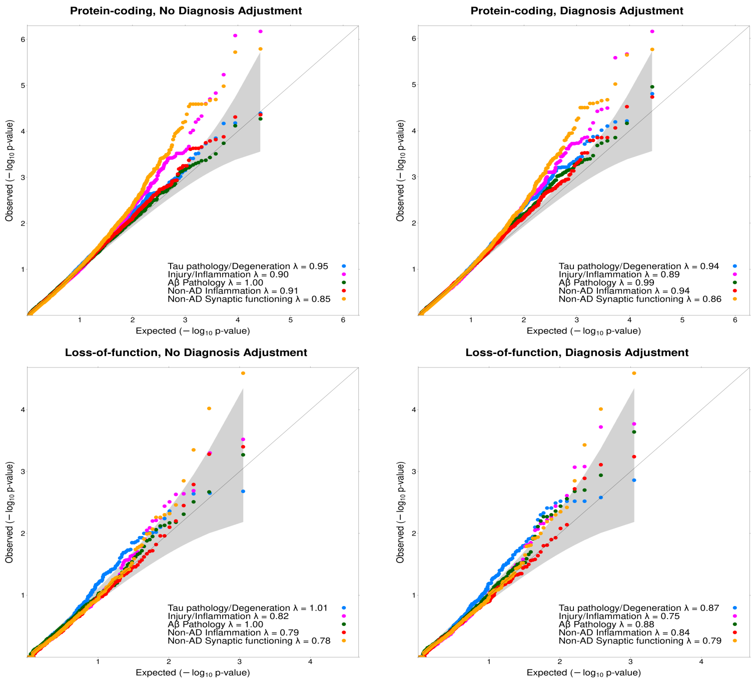

**Figure S2: Manhattan plot of the exome-wide rare variant analyses (loss-of-function).** Results from the exome-wide rare variant (MAF < 1%) analyses of five CSF biomarker principal components (PC) (n=480). Each plot displays a different PC as outcome. X-axis represents each gene (loss-of-function variants) and the y-axis the p-value obtained from gene-based SKAT-O tests on a $-\log_{10}$ scale. All analyses were adjusted for sex, age and genetic ancestry. Blue points represent p-values additionally adjusted for diagnosis. Red line indicates exome-wide significance threshold ($p=1.9*10^{-4}$). Yellow line indicates suggestive threshold ($p=3.7*10^{-3}$).

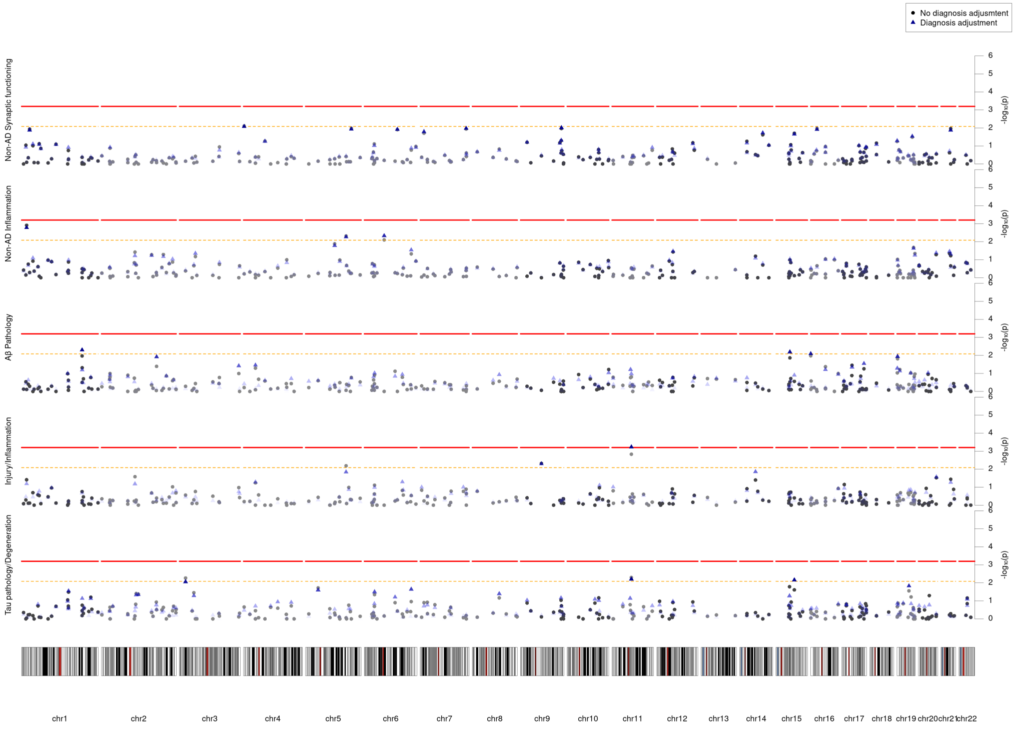
